# Simulation Models for Bladder Cancer: A Scoping Review

**DOI:** 10.1101/2025.03.17.25324125

**Authors:** Stavroula A. Chrysanthopoulou, Timothy Hedspeth, Dana Antinozzi, Andrew W. Huang, Yullia Sereda, Hawre Jalal, Thomas A. Trikalinos, John B Wong, Stella K. Kang

## Abstract

**Objectives:** The study identifies and summarizes information from manuscripts using simulation models for Bladder Cancer (BCA) research.

**Methods:** We conducted and presented results of a systematic literature search of Medline, Web of Science, and Google scholar, following the PRISMA guidelines for scoping reviews. We summarized extracted key components of the methodology, data sources, and software used for the development of simulation models and classify eligible articles in terms of the study objectives and conclusions.

**Results:** The 97 identified modeling studies simulating aspects of BCA included models that (1) describe the biological process of carcinogenesis and tumor progression (mostly compartmental models); (2) examine the impact of screening protocols and interventions on disease progression and prognosis (mostly microsimulation models); and (3) assess the cost-effectiveness of BCA treatment and control strategies (cohort-based simulation models or simpler decision tree structures). The scope, objectives, and conclusions of these studies varied substantially. Most focused on evaluating treatments, mostly for non-muscle invasive bladder cancer, with some examining BCA screening and surveillance. Their objectives, methods, and analyses were inconsistently and often incompletely reported.

**Conclusions:** Simulation models in bladder cancer examine questions that span the range from tumor kinetics to cost effectiveness of tumor management, but shortcomings in their reporting hinder assessments of their applicability and methodological rigor, severely limiting their practical usefulness.

**Highlight statements:**

- We assessed the available landscape of simulation modeling for health decision making in BCA research.
- Shortcomings in the reporting of this research severely limit their practical usefulness.
- Future population modeling should assess BCA screening and surveillance.

**Strengths:**

- This is the first, to our knowledge, systematic appraisal of simulation models in bladder cancer. Simulation modeling will be a key technology to assess the utility of highly promising novel diagnostics and treatments, while evidence accumulates.
- The described variation in the objectives, methodological rigor, and reporting of models’ development, validation, and analysis likely generalize to other disease areas.

**Limitations:**

- This descriptive compendium does not explicitly compare the results of different models between them or with observed data.

## Introduction

Bladder cancer has the 6^th^ highest cancer incidence in the United States, more commonly occurring in men (4^th^ highest incidence) than women (11^th^ highest incidence) [1]. Non-muscle invasive bladder cancer (NMIBC) comprises about 70% of all newly diagnosed bladder cancers and includes tumors at stages Ta, T1, and carcinoma in situ, with varied risks of recurrence and prognosis depending upon patient and lesion characteristics. The median age of diagnosis of bladder cancer is 73 and of dying from bladder cancer is 79 years with an overall 5-year relative survival of around 78%[1]. Despite being diagnosed most often as non-muscle-invasive disease (70%), high-risk NMIBC as defined using the American Urological Association has a 45% annual rate of progression to muscle invasive or metastatic disease[2]. Urinary bladder cancer prevention and control would benefit from having an effective screening tests and more personalized safe and effective management strategies.

Current recommendations for bladder cancer (BCA) intervention and surveillance are based on the available evidence and stakeholder opinions. All international guidelines on the management of NMIBC recommend cystoscopy for surveillance based on the patient’s risk at varying intervals [56]. BCA surveillance involves cystoscopy that is associated with patient discomfort, risk of complications, and high costs. In addition, the frequent recurrence and need for further intravesical chemotherapy when disease remains NMIBC, or other more invasive therapies when disease progresses, leads to cumulatively high costs with many possible treatment options. In order to assess the potential effectiveness of management pathways, clinical trials are not often feasible due to cost, the time required to find mortality differences, and the plethora of options that could be explored.

Simulation modeling can fundamentally advance research in cancer prevention and control by identifying the management strategies that provide the best balance of control and harms. Simulation models have been developed and published to describe the dynamics of BCA, compare treatment strategies using projected outcomes, and evaluate interventions under different scenarios. However, it remains unclear how existing models capture facets of risk factors, heterogeneity in tumor aggressiveness, and clinical practice patterns, among other influential assumptions. Evidence-synthesis methods systematically combine information from different sources to inform models that address management questions including screening frequency, comparative effectiveness, uncertainty, costs, and trade-offs between benefits and risks of the main procedures are weighed. The main objective of this study was to search the literature systematically for original studies using simulation models and characterize the most common objectives, main practices in their development, and applicability to understanding BCA outcomes. We also sought to identify important gaps in the existing literature on bladder cancer simulation modeling studies.

To our knowledge, this is the first literature review of relevant models for BCA. This review will clarify the terminology and nomenclature, summarize common practices, describe models’ data sources, and identify gaps in the literature with areas for improvement. This study was motivated by our work as part of the CISNET bladder cancer incubator site, for developing three comprehensive, microsimulation models for BCA (COBRAS, Kystis, and SCOUT).

## Methods

We searched PubMed, Web of Science, and Google Scholar to identify original studies that involved mathematical models to conduct simulation-based analyses [3, 4] on important questions in bladder cancer research. We applied a scoping review methodology for summarizing the results from this review, given the broad range of potential focus areas, modeling methodologies, applications, and conclusions possible in these studies. This study adheres to the PRISMA-ScR guidelines for scoping reviews [5].

### Search strategy

To identify relevant articles, we specified keywords to apply to a broad search strategy reflecting the widely variable terminology in the field. The search algorithm included: “Bladder cancer” AND “model” AND any of the terms in a combined ‘OR’ statement: “Simulation” OR “Microsimulation” OR “State Transition” OR “Dynamic Transition” OR “Multistate” OR “Monte Carlo Simulation” OR “Risk Prediction” OR “Cohort” OR “Cohort-based” OR “Population” OR “Population-based”.

### Eligibility Criteria

We included manuscripts describing development and implementation of complex mathematical models used for simulation of disease development and progression in bladder cancer. These studies frequently address research questions related to mechanisms of tumor development or progression, as well as evaluation of healthcare interventions (treatment, screening, and surveillance) for BCA. We included articles written in English, without restricting scope of study purpose, published until May 2022. Articles referring to the same model for BCA were included in the analysis if they pertained to applications to different target populations, model extensions to account for different facets of the disease, or novel study objective (answering different research questions). We excluded articles that reflected meta-analyses, literature reviews, statistical modeling of empirical datasets, editorial comments, or letters to the editor along with gray literature such as unpublished reports, conference presentations, and meeting proceedings.

After deduplicating redundant reports, two reviewers (TH and DA) independently screened titles and abstracts to identify potentially relevant papers, which were then examined in full text. All discrepancies were discussed with and by a third reviewer (SC).

### Data Extraction

We extracted information in structured electronic forms on (1) model technical details (including model type, structure, development, performance), (2) study objectives, (3) comparison of results for models with similar objectives, and (4) reporting of modeling questions, objectives, and methods. Briefly, we systematically categorized models based on their key features including model structure, model outputs, simulation approach, underlying population, and time consideration (simulating continuous vs discrete time steps). We also collected technical features: calibration and validation methods, evaluation of their predictive accuracy, and uncertainty propagation. We extracted the key research questions that these models aimed to answer related to BCA risk, progression, intervention, and surveillance captured the aim for use of simulation models for questions pertaining to public health and clinical decision making.

We reviewed the key objectives and identified common or contradictory findings from the eligible studies. We also evaluated the reporting practices with specific gaps in nomenclature, terminology, technical components, and model transparency.

We used EndNote reference software for management of identified manuscripts (i.e., de-duplication, storage), Microsoft Excel for data extraction, and the R statistical software [6] for summarizing findings of this scoping review.

## Results

Figure 1 shows the literature flow, including reasons for and numbers of excluded papers. Of the 4871 articles returned by the searches, 2570 were reviewed in full text and 97 were included in the scoping review.

**Figure 1.**
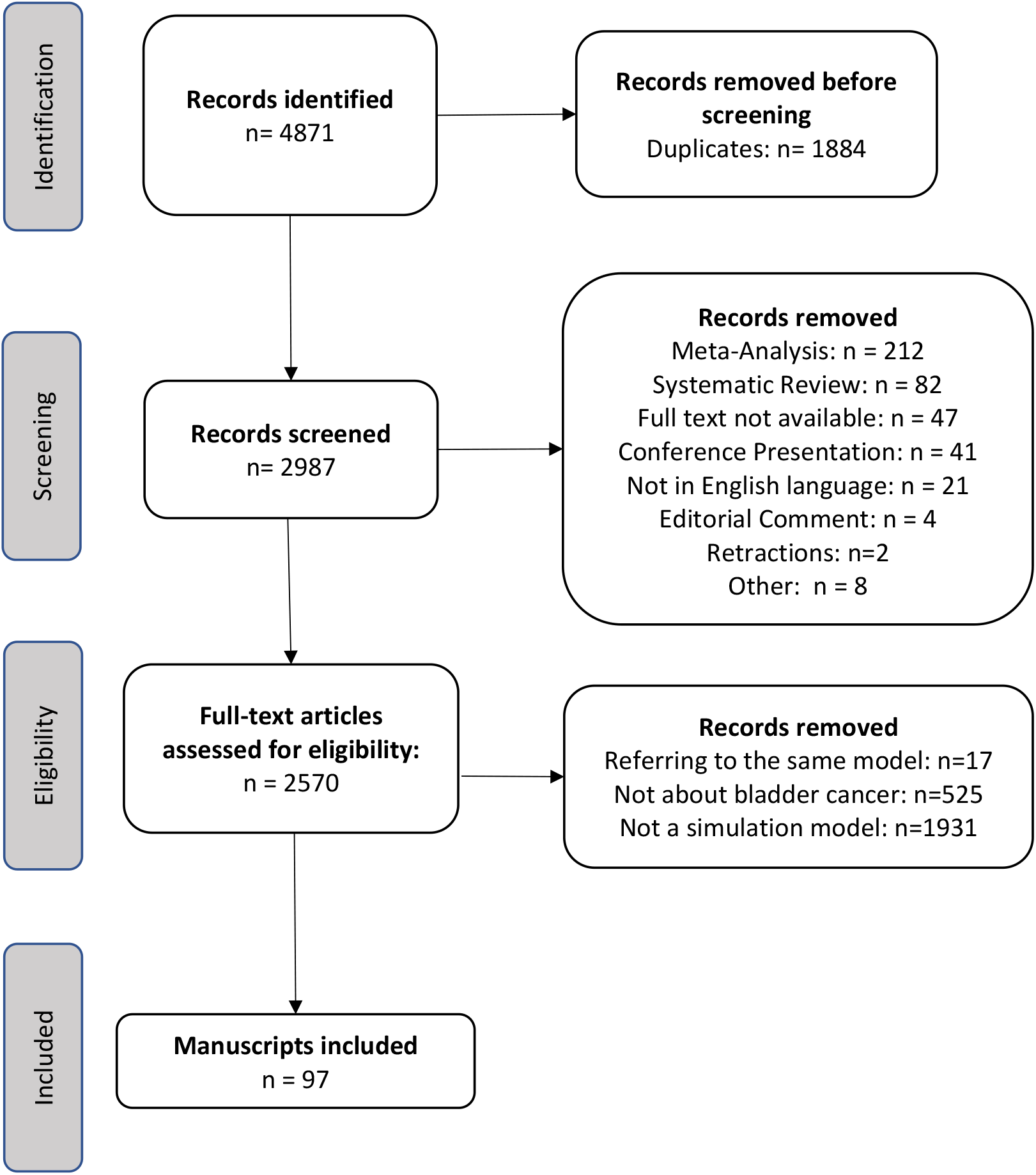
PRISMA flow diagram of the scoping review of simulation models for bladder cancer.

### Publication Journals

The number of published simulation models for BCA increased over time (Figure 2). Table 1 shows the Web Of Science categorization of the journals in which eligible papers were published [7]. Most manuscripts were published in Clinical Medicine and Computational Biology journals, with others published in in Applied Mathematics, Engineering, Health Services Research, or multidisciplinary fields. Most studies published in medical journals focused on urologic oncology or oncology. Some studies modeling carcinogenesis and disease progression processes and tumor kinetics were also published in Computational Biology, Statistics, and Applied Mathematics journals.

**Table 1.**
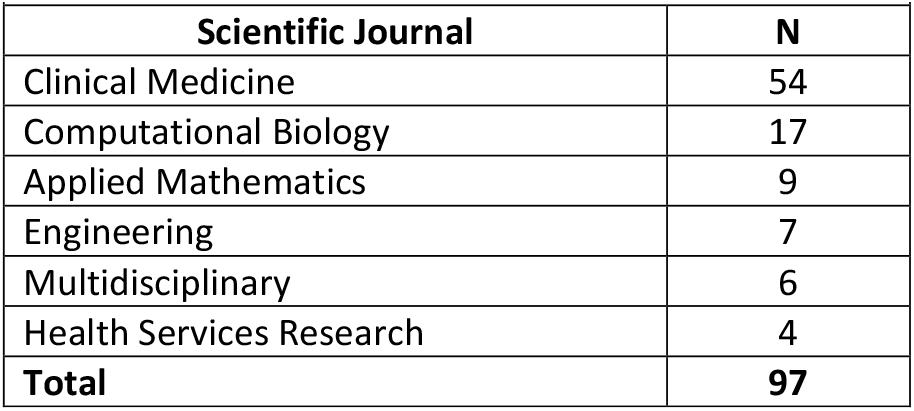
Published articles of studies using simulation models for bladder cancer by scientific journal JRC classification [7].

| **Scientific Journal** | **N** |
| --- | --- |
| Clinical Medicine | 54 |
| Computational Biology | 17 |
| Applied Mathematics | 9 |
| Engineering | 7 |
| Multidisciplinary | 6 |
| Health Services Research | 4 |
| **Total** | **97** |

**Figure 2.**
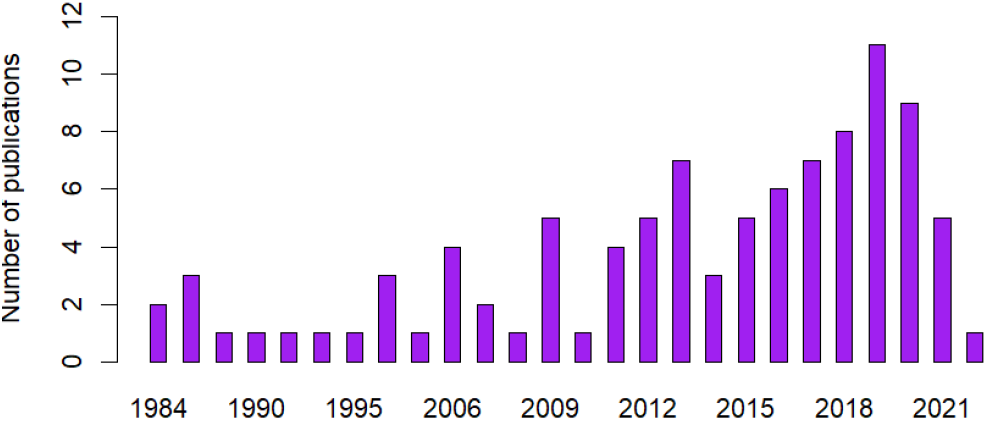
Publication trend of simulation modeling studies for bladder cancer over time with the exception of the COVID-19 pandemic years.

### Model Types

In terms of their structure and simulation approach, we classified included models into four broad categories.

1. *Decision trees* (n=17) were used for cost-effectiveness analysis (CEA, e.g., cost-utility analysis (CUA), etc.) that aimed to comparing alternative bladder interventions using the incremental cost-effectiveness ratio (ICER) [8-24].
2. *Compartmental models* (n=37), were the most commonly used type for describing the dynamics of BCA with emphasis on the biological process of cancer development and progression [25-61].
3. *Population-/cohort-based simulations* (n=13) were used to assess port NMIBC treatment surveillance and management of bladder cancer. [97-109]
4. individual-based microsimulation models (MSMs) (n=28) were used to describe the natural history of bladder cancer and to evaluate screening protocols and treatments on related outcomes of interest [62-89].

### Population Characteristics

We identified three types of studies: *in vivo, in vitro*, and *in silico*.[97]

The majority (n=86) of the *studies* involved modeling of human subjects vs a few biological studies on mice, dogs, and pigs. Of the studies in human populations, 57 specified the country of the reference population, of which most were focused on the US (n=36) or European countries (13). (Section S.4 – appendix). Three studies compared data from the US with a European country and one compared policies between US, UK, Australia, and Canada.

### Data sources

Some of the most commonly used data sources for fitting and calibrating simulation models for BCA were from the Surveillance, Epidemiology, and End Results (SEER) database[98], European Organisation for Research and Treatment of Cancer (EORTC), National Cancer Database (2003–2012) [99], and CDC (Centers for Disease Control and Prevention). Some used an institutional (University of Michigan Medical School) bladder cancer database or the Healthcare Cost and Utilization Project State Inpatient Databases for CEA analysis.

### Overview of Bladder Cancer Characteristics in Modeling Studies

Simulation modeling studies examined non-muscle invasive bladder Cancer (NMIBC) (n=46), muscle invasive bladder cancer (MIBC) (n=31), and metastatic BCA (n=18). Many studies simulate BCA development and progression without specifying a particular histologic subtype of BCA (i.e., mainly in compartmental models, most of which describe the carcinogenesis process without a specific reference to any BCA type. Section 7 in the appendix provides details on studies that do not focus on a specific type of cancer). Studies on metastatic BCA often use cohort-based simulation models, while decision trees, microsimulation, and cohort-based simulation models have been used for NMIBC and MIBC. (Section S.5 – appendix)

The treatments evaluated with simulation models include Bacillus Calmette-Guerin (BCG) immunotherapy, transurethral resection of bladder tumor (TURBT), mytomycin C (MMC), various chemotherapy regimens, as well as surgical treatment with cystectomy (Section S.6 – appendix). Key outcomes usually included BCA diagnosis, recurrence, and mortality. Several models also made specific assumptions about tumor growth, as detailed below. The effectiveness of BCA interventions was usually measured in quality-adjusted life years (QUALY, QALY) or quality-adjusted life expectancy (QALE), cost, and incremental cost-effectiveness ratio (ICER), specific to bladder cancer type and stage.

### Modeling Objectives

The included studies’ objectives fell into four broad research areas, namely (1) describing tumorigenesis and tumor kinetics, and comparing the effectiveness of (2) BCA treatments, (3) screening, and (4) surveillance. A minority of studies examined miscellaneous objectives (e.g., impact of arsenic exposure) which are not summarized in the text (see Appendix S.10).

#### 1. Tumorigenesis and tumor kinetics

Several models, mostly compartmental and MSMs, modeled biological processes and cellular dynamics of BCA (initiation and progression). Examples include modeling of the Mitogen-Activated Protein Kinase (MAPK) signaling network; the regulatory network around E2F (a family of transcription factors for which disregulation has been associated with cancer progression, chemoresistance, invasiveness, and metastasis), Epidermal Growth Factor Receptor (EGFR) over-expression and Fibroblast Growth Factor Receptor 3 (FGFR3) activating mutations [70, 86] and the impact of exposure to nongenotoxic compounds (e.g., sodium saccharin) and other factors on carcinogenesis [69].

#### 2. Comparative and cost effectiveness of treatments

Several models assessed the effectiveness or cost-effectiveness of the initial management of NMIBC by comparing various protocols for administering BCG alone or in combination with other treatments such as interleukin-2 (IL-2), thermotherapy, hexaminolevulinate hydrochloride-guided blue-light flexible cystoscopy (HAL BLFC). The assessed strategies varied widely across publications. They included comparisons across different regimens; size and timing of doses; use of blue light cystoscopy; use of TURBT, fulguration, and laser ablation, in all NMIBC or by patient-specific biological, morphological, environmental, and clinical characteristics.

The effectiveness and cost-effectiveness of MIBC management strategies was examined in several publications. Models compared open and robot assisted radical cystectomy, immediate cystectomy, conservative therapy followed by delayed cystectomy, and trimodal therapy.

An additional 4 studies compared pembrolizumab, avelumab, and atezolizumab as second-line treatments for metastatic disease [21, 22, 64, 100].

#### 3. Comparative and cost effectiveness of screening

Multiple simulation studies (31 in total; Section S.4 – appendix) evaluated the potential and cost-effectiveness of diagnostic tests for BCA screening, including (renal) ultrasound, multiphase computerized tomography, cystoscopy, voided urine cytology, quantitative fluorescence image analysis (QFIA), routine urinalysis for microhematuria, home dipstick testing, and novel, urine-based tumor markers, such as BladderChek (NMP22). Variations of cystoscopy such as hexaminolevulinate hydrochloride-guided blue-light flexible cystoscopy (HAL BLFC) and white light cystoscopy (WLC) have also been explored as screening alternatives.

#### 4. Cost-effectiveness of surveillance

MSMs have been used to also evaluate the cost-effectiveness of different surveillance schedules for NMIBC in older adults. They examined surveillance based on cystoscopy only or aided by biomarkers; general versus age-specific surveillance recommendations, as well as recommendations based on histologic grade, tumor stage, and number of prior tumors as risk-stratification factors.

### Technical Details

Unfortunately, incomplete reporting practices made it difficult or impossible to evaluate the performance and validity of the models and to reproduce the study results. Often, little or no information was provided about important aspects of model development. Few studies included details about model calibration and validation approaches. Several studies did present some type of sensitivity analysis for evaluating the impact of different sources of uncertainty (mainly parameter uncertainty) (Section S.8 – appendix).

Most of the studies provided graphical representations of the model structure (distinct health states, transition rules, decision alternatives, etc.). The most broadly used software for the development of simulation models for BCA has been TreeAge (mainly for decision trees used in cost-effectiveness analyses). MATLAB, C++, and R have been used especially for models of a more complicated structure (compartmental or MSMs). In most cases detailed description of the model structure and assumptions remains lacking, and the source code of the model is unavailable. (Section S.9 – appendix)

## Discussion

This scoping review of available literature involving simulation modeling in BCA research finds increasing publications trends in their use to answer questions related to BCA with relevant studies most commonly published in medical rather than statistical, computer science, or other technical scientific journals. The considerable amount of effort dedicated to the development and use of simulation models in BCA research demonstrates the utility and need for simulation models to help synthesize relevant information from the literature and derive helpful conclusions based on available data and simulated trends and findings.

The main objectives of eligible simulation studies can be classified into four broad groups around carcinogenesis, treatment, screening, and surveillance. However, there remains large variability in objectives and hence conclusions from these studies. In addition, one of the major issues when searching the literature is the lack of consensus in the simulation model terminology and nomenclature. This substantial heterogeneity in both scope and methodologies of relevant studies hinder a comprehensive synthesis and comparative understanding of the available research. It renders the classification of relevant work into meaningful categories rather difficult.

Overall low quality of reporting practices also indicated a need for determining specific guidelines to standardize model terminology and nomenclature, to facilitate sharing of information, harmonization, and even collaboration between modelers and clinical researchers with interest in these tools for investigation.

Considerable research has focused on the comparative effectiveness of treatment and screening protocols for BCA, indicating the interest and potential value to advanced evidence-synthesis techniques for informed decision making in this area. Most BCA models currently address US population outcomes with results focused on trends and findings for that population. Comparatively, there are fewer publications for metastatic BCA.

Unfortunately, the apparent lack of description in many studies of systematic calibration and validation of those models to real data may decrease the face validity acceptance of the models, increase the risk of bias, or even worst, result in contradictory findings across decision analyses on clinical practices. Code is usually not publicly available and reporting practices from these studies remain incomplete, making the research non-reproducible research due to lack of model transparency (assumptions, structure, etc.), and hence difficulty in evaluating and comparing relevant results.

Although simulation models have been widely used to explore important facets of the disease, there remains a lack of large-scale, complex predictive models synthesizing best available data from multiple sources to describe the dynamic evolution of bladder cancer with the detail, sophistication, and nuance necessary to evaluate alternative control and prevention interventions and key population groups for health policy decisions in bladder cancer prevention, screening, surveillance, and treatment in a comprehensive and trustworthy manner.

## Data Availability

All data produced in the present work are contained in the manuscript.

## Acknowledgments

This work has been supported by the by the National Cancer Institute (NCI) of the National Institutes of Health (NIH) under award numbers U01CA265750 and R01CA262375.

Patients and the public were not involved in the design, conduct or reporting of this review of previously published data.

