## Supplemental Material for "Simulation Models for Bladder Cancer: A Scoping Review"

### S.1 Abbreviations

| Table 1. Abbreviations | | |
|---|---|---|
| **Abbreviation** | **Description** | **Notes** |
| AC | adjuvant chemotherapy | |
| ACS | American cancer society | |
| AMH | asymptomatic microscopic hematuria | |
| AS | Arsenic | |
| AUA | American Urological Association | |
| BCG | Bacillus Calmette-Guerin | intravesical therapy |
| BIM | Budget Impact Model | |
| BLC | blue light cystoscopy | |
| BM model | Bunimovich-Medrazitsky mathematical model | |
| CA | Cellular Automata | |
| CBC | Cisplatin-based Chemotherapy | |
| CDC | Centers for Disease Control and Prevention | |
| CH | Chitosan | |
| C-HT | Chemo-Hyperthermia Treatment | |
| CIS | Carcinoma in situ | |
| CSC | Cancer Stem Cell | |
| CT | computed tomography | |
| CTU | computerized tomography urogram | |
| CUA | Canadian Urological Association | |
| CUA | Cost-Utility Analysis | |
| DT | Decision Tree | |
| EAU | European Association of Urology | |
| EBRT | External Beam Radiation Treatment | |
| EGFR | Epidermal Growth Factor Receptor | |
| EORTC | European Organisation for Research and Treatment of Cancer | https://www.eortc.org/ |
| ERC | Early Radical Cystectomy | |
| ERR | Excess Relative Risk | |
| ETS | Environmental Tobacco Smoke | |
| FANFT | N-[4-(5-nitro-2-furyl)-2-thiazolyl]formamide | |
| GI | Glycemic index | |
| GNR | Gold nanorods | |
| GNR-PTT | Gold nanorods assisted photothermal therapy | |
| GU | Genitourinary | |
| HAL | Hexaminolevulinate | |
| HAL BLFC | Hexaminolevulinate hydrochloride-guided blue-light flexible cystoscopy | |
| HDR-BT | High-Dose-Rate Brachy Therapy | |
| HIPEC | Hyperthermic Intraperitoneal Chemotherapy | |
| HRBC | High-risk NMIBC | |
| IC | Inpatient Cystodiathermy | |
| ICCD | Incremental Cost per Cancer Detected | |
| ICER | Incremental Cost-Effectiveness Ratio | |
| ICI | Immune Checkpoint Inhibitors | |
| IL-2 | Interleukin-2 | |

| Table 1. Abbreviations | | |
|---|---|---|
| **Abbreviation** | **Description** | **Notes** |
| ImmPort | Immunology Database and Analysis Portal | |
| IRBC | Intermediate-risk NMIBC | |
| IVIC | intravesical instillation of chemotherapy | |
| LE | Life Expectancy | |
| LHS | Latin Hypercube Sampling | |
| LMR | Lymphocyte–monocyte ratio | |
| LRDF | Low-risk disease free | |
| LYGS | Life-year gains | |
| MAPK | Mitogen-Activated Protein Kinase | |
| MCM | Medical Countermeasures | |
| MDACC | MD Anderson Cancer Center | Houston, Texas |
| MetS | Metabolic syndrome | |
| MGR | Multi-Generation Register | Sweeden data base |
| MIBC | muscle-invasive bladder cancer | |
| MMC | mitomycin C | |
| mBC | metastatic Bladder Cancer | |
| mUC | metastatic Urothelial Cancer | |
| MVAC | methotrexate, vinblastine, doxorubicin and cisplatin | Type of chemo regiment |
| NCDB | National Cancer Database (US) | From ACS and American College of surgeons |
| NHI | National Health insurance program | Data base from Taiwan |
| NMACB | non-metastatic primary adenocarcinoma of the bladder | |
| NMIBC | Non–muscle-invasive bladder cancer | also called superficial |
| NU | nephroureterectomy | |
| OBF | Office-based Bladder Tumor Fulguration | |
| OC ( ORC) | Open (Radical) Cystectomy | |
| OS | Overall Survival | |
| OR | Operating Room | |
| OS | Overall Survival | |
| PBPK | Physiologically-Based Pharmacokinetic Model | |
| PC | Partial Cystectomy | |
| PC (trt) | polycarbophil | Chemotherapy |
| PD | Pharmacodynamics | |
| PD-1 | programmed cell death protein 1 | |
| PDD | Photodynamic Diagnosis | |
| PDE | Partial Differential Equations | |
| PFS | Progression-free survival | |
| PK | Pharmacokinetics | |
| PLCO | Prostate, Lung, Colorectal and Ovarian Cancer Screening Trial | |
| POI | Postoperative ileus | Complication after RC |
| PTT | Photothermal Therapy | |
| QALE | Quality Adjusted Life Expectancy | |
| QALY/QUALY | Quality Adjusted Life Years | |
| QFIA | quantitative fluorescence image analysis | |
| RARC | Robot-assisted Radical Cystectomy | |
| RC | Radical cystectomy | |
| REID | Risk of Exposure Induced Death | |
| RNU | radical nephroureterectomy | |

| **Table 1. Abbreviations** | | |
|---|---|---|
| **Abbreviation** | **Description** | **Notes** |
| SPVF | Singularly Perturbed Vector Field | |
| T1 | stage of bladder. cancer where the wall is not involved | also superficial |
| Ta | noninvasive papillary carcinoma | |
| TCC | Transitional Cell Carcinoma | |
| TIS | transitional cell carcinoma in-situ | also considered superficial |
| TNM | TNM staging system: A system to describe the amount and spread of cancer in a patient's body, using TNM.<br>T describes the size of the tumor and any spread of cancer into nearby tissue; N describes spread of cancer to nearby lymph nodes; and M describes metastasis (spread of cancer to other parts of the body). This system was created and is updated by the American Joint Committee on Cancer (AJCC) and the International Union Against Cancer (UICC). The TNM staging system is used to describe most types of cancer. | Also called AJCC staging system. |
| TUR | treated surgically by transurethral resection (TUR) | |
| TURB/TURBT | transurethral resection of bladder tumor | |
| UC | urotherial cancer | Also considered superficial |
| UTUC | Upper Tract Urothelial carcinoma | |
| WLC | white light cystoscopy | |
| WTP | Willingness-to-pay (in CEA) | |

### *S.2 Scientific Journals publishing studies based on simulation modeling*

Table 2. Published articles of studies using simulation models for bladder cancer by scientific journal.

| **Scientific Journal** | **n(%)** |
|---|---|
| BJU International | 3 (3.1) |
| Bulletin of Mathematical Biology | 2 (2.1) |
| Cancer Research | 2 (2.1) |
| Clinical Genitourinary Cancer | 2 (2.1) |
| Computers in Biology and Medicine | 2 (2.1) |
| European Urology Focus | 2 (2.1) |
| Journal of Theoretical Biology | 2 (2.1) |
| Journal of Toxicology and Environmental Health-Part B-Critical Reviews | 2 (2.1) |
| Mathematical Biosciences and Engineering | 4 (4.1) |
| Mathematical Methods in the Applied Sciences | 2 (2.1) |
| Medical Decision Making | 3 (3.1) |
| Medical Physics | 2 (2.1) |
| PLOS One | 2 (2.1) |
| Physics in Medicine & Biology | 2 (2.1) |
| Risk Analysis | 2 (2.1) |
| Statistics in Medicine | 2 (2.1) |
| The Journal of Urology | 8 (8.2) |
| Urologic Oncology: Seminars and Original Investigations | 4 (4.1) |
| Urology | 3 (3.1) |
| Other | 46 (47.4) |
| **Total** | **97** |

Summaries are presented as frequencies (n) and percentages (%).

### *S.3 Model Nomenclature*

There is not a consensus on the model nomenclature, thus making it hard to identify and classify simulation models of similar nature, structure, and specifications. **Table 3** shows the names that eligible manuscripts use when referring to each one of the four main types of simulation models identified in this systematic review.

**Table 3. Simulation model nomenclature.**

| **Model Type** | **Model name in the manuscript** |
|---|---|
| Decision tree | Decision analytic (or analytical or analysis) model |
| | Decision tree |
| | Markov model |
| | Markov state transition model |
| | Simulation model |
| Markov Cohort-/Population- based | Decision analytic model |
| | State transition model |
| | Markov (cohort) state transition model |
| | Markov decision analysis model |
| | Multi-state Markov model |
| | Random walk cancer model |
| Microsimulation | Computational model |
| | Computational network modeling |
| | Discrete event simulation |
| | Homogeneous, time-continuous Markov model |
| | Logical model |
| | Markov model |
| | Markov state transition model |
| | Mathematical model |
| | Microsimulation |
| | Monte Carlo simulations |
| | Multistate model |
| | Numerical finite-difference time-domain (fdtd) model |
| | Partially observable Markov model |
| | Simulation model |
| | State transition model |
| Compartmental | Compartmental |
| | Kinematic model |
| | Logic-based model |
| | Mathematical model |
| | Mathematical model using ordinary differential equations |
| | Monte Carlo simulation algorithm |
| | Numerical integration over discrete time intervals |
| | Numerical simulations |
| | Population four-compartment model |
| | Risk model |
| | Stochastic model |

### *S.4 Simulated population and key outcomes of interest*

**Table 4. Key features (input and output) of simulation models for BCA.**

| Model input | | | Model Output | |
|---|---|---|---|---|
| **Description** | **n** | | **Description** | **n** |
| **Subjects** | | | **Diagnosis/Screening** | |
| Human | 86 | | Yes | 31 |
| Mouse | 3 | | No | 67 |
| Rat | 2 | | | |
| Dog | 2 | | **Mortality** | |
| Pig | 1 | | Yes | 23 |
| Human and Mouse | 3 | | No | 74 |
| **Country** | | | **Recurrence** | |
| Unknown | 21 | | Yes | 39 |
| N/A | 19 | | No | 58 |
| United States | 36 | | | |
| United Kingdom | 4 | | **Tumor growth** | |
| Spain | 3 | | Yes** | 27 |
| Other | 14 | | No | 70 |

** Specific distribution/assumption

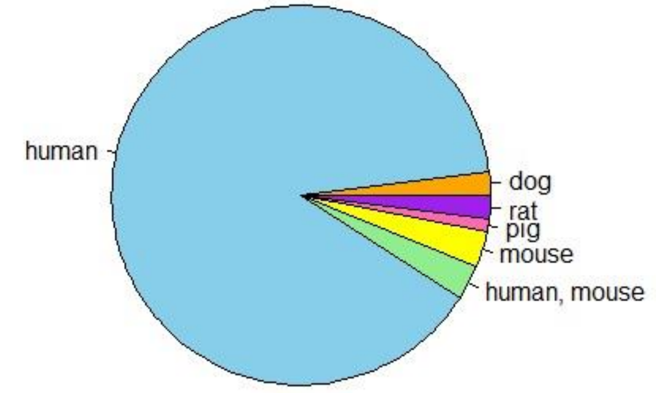

Figure 1. Simulated (reference) population in eligible studies.

Table 5. Reference human population explored in the simulation studies.

| **Country** | **n** |
|---|---|
| N/A | 19 |
| Unknown | 21 |
| United States (US) | 37 |
| United Kingdom (UK) | 4 |
| Spain | 3 |
| Canada | 2 |
| Netherlands | 2 |
| Sweden | 2 |
| Denmark | 1 |
| France | 1 |
| Taiwan | 1 |
| Tanzania | 1 |
| US, EORTC | 1 |
| US, France | 1 |
| US, UK | 1 |
| US, UK, Australia, Canada | 1 |

Table 6. Simulated population by model type.

| | Species | | | | | | |
|---|---|---|---|---|---|---|---|
| Model Type | Human | Human, mouse | Mouse | Dog | Rat | Pig | Total |
| Compartmental | 32 | 2 | 1 | 1 | 0 | 1 | 37 |
| Microsimulation | 23 | 1 | 2 | 0 | 2 | 0 | 28 |
| Decision Tree | 17 | 0 | 0 | 0 | 0 | 0 | 17 |
| Cohort-based simulation | 13 | 0 | 0 | 0 | 0 | 0 | 13 |
| Decision Tree and Microsimulation | 1 | 0 | 0 | 0 | 0 | 0 | 1 |
| Other | 0 | 0 | 0 | 1 | 0 | 0 | 1 |
| Total | 86 | 3 | 3 | 2 | 2 | 1 | 97 |

### *S.5 Type of BCA explored with simulation modeling*

The most common types of bladder cancer explored with simulation models are the NMIBC and MIBC.

**Table 7. Type of bladder cancer explored.**

| Bladder Cancer Type | N |
|---|---|
| General | 35 |
| Nmibc | 25 |
| nmibc, mibc, metastatic | 11 |
| Mibc | 9 |
| nmibc, mibc | 9 |
| Metastatic | 5 |
| nmibc, metastatic | 1 |
| mibc, metastatic | 1 |
| Upper track urothelial carcinoma | 1 |
| **Total** | **97** |

**Table 8.** Type of bladder cancer examined by type of simulation model (details).

| Model Type | Cancer Type | Freq | % |
|---|---|---|---|
| Cohort-based | General | 4 | 4.12 |
| | Metastatic, mibc, nmibc | 3 | 3.09 |
| | nmicb | 2 | 2.06 |
| | Metastatic | 1 | 2.06 |
| | Metastatic, mibc | 1 | 1.03 |
| | Metastatic, nmibc | 1 | 1.03 |
| | Nmibc,micb | 1 | 1.03 |
| Compartmental | General | 19 | 19.6 |
| | nmibc | 14 | 14.4 |
| | mibc | 2 | 2.06 |
| | Mibc, nmibc | 2 | 2.06 |
| Decision Tree | General | 5 | 5.15 |
| | nmibc | 5 | 5.15 |
| | mibc | 3 | 3.09 |
| | Metastatic, mibc, nmibc | 2 | 2.06 |
| | Metastatic | 2 | 2.06 |
| Microsimulation | General | 6 | 6.19 |
| | mibc, nmibc | 6 | 6.19 |
| | Metastatic, mibc, nmibc | 5 | 5.15 |
| | Micb | 4 | 4.12 |
| | Nmicb | 4 | 4.12 |
| | Metastatic | 2 | 2.04 |
| | Upper tract urothelial carcinoma | 1 | 1.03 |

### *S.6 Type of BCA treatments compared with simulation modeling*

| Table 9. Number of articles mentioning evaluation of the specific BCA treatments. | |
|---|---|
| **BC treatments** | **N** |
| adjuvant chemotherapy, rc | 1 |
| atezolizumab, chemotherapy | 1 |
| Bcg | 12 |
| bcg, chemotherapy, cystectomy, turbt | 1 |
| bcg, chemotherapy, interferon, mmc, rc | 1 |
| bcg, chemotherapy, mmc | 1 |
| bcg, chemotherapy, rc | 2 |
| bcg, cystectomy, mmc, neoadjuvant chemotherapy, trimodal therapy, turbt | 1 |
| bcg, il-2 | 4 |
| bcg, mmc, rc, turbt | 1 |
| bcg, mmc, turbt | 1 |
| bcg, pc, turbt | 1 |
| bcg, rc | 1 |
| chemotherapy | 4 |
| chemotherapy, cystectomy | 1 |
| chemotherapy, cystectomy, fulguration, turbt | 1 |
| chemotherapy, cystectomy, mmc, turbt | 1 |
| chemotherapy, fulguration, turbt | 1 |
| chemotherapy, immunotherapy | 3 |
| chemotherapy, irradation, rc, tur | 1 |
| chemotherapy, nephroureterectomy | 1 |
| cystectomy | 1 |
| cystectomy, irradation, tur | 1 |
| cystectomy, neoadjuvant chemotherapy | 1 |
| cystectomy, neoadjuvant chemotherapy, rc, trimodal therapy | 1 |
| cystodiathermy, laser ablation | 1 |
| fulguration, turbt | 1 |
| gnr-ptt | 1 |
| hyperthermia | 2 |
| N/A (initial treatment) | 1 |
| Mmc | 1 |
| mmc, thermotherapy | 2 |
| nanoparticle-assisted photothermal therapy | 1 |
| neoadjuvant chemotherapy | 1 |
| neoadjuvant chemotherapy, orc, rarc | 1 |
| neoadjuvant chemotherapy, rc, tmt | 1 |
| nephroureterectomy, rc, tur | 1 |
| orc, rarc | 1 |
| perioperative chemotherapy, rc, turbt | 1 |
| radical cystectomy | 1 |
| rc | 2 |
| turbt | 1 |
| turbt, cystectomy | 1 |

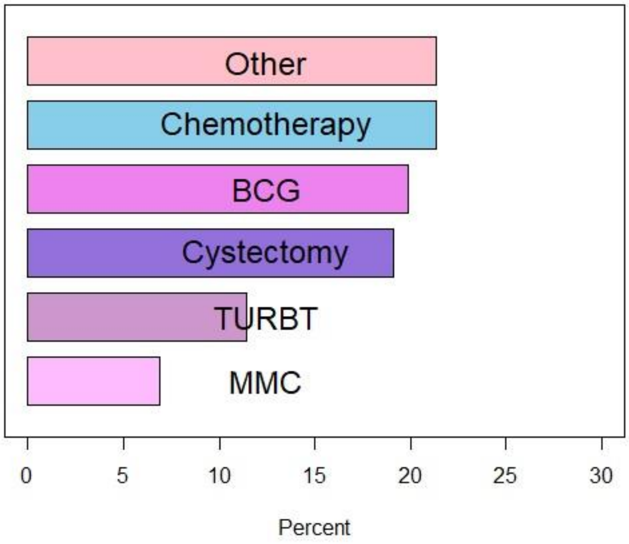

Figure 2. Type of treatment included in the simulated model

**Table 10. Type of BC treatment evaluated with simulation modeling**

| Treatment* | N |
|---|---|
| Bacillus Calmette-Guerin (BCG) | 26 |
| Chemotherapy | 28 |
| Cystectomy | 25 |
| Mitomycin C | 9 |
| Transurethral Resection of Bladder Tumor (TURBT) | 15 |
| Other | 28 |

* Note that the total is not n=97 (total number of eligible studies) as there are several studies examining the performance of multiple therapies.

### *S.7 Details from studies not focusing/mentioning a specific type of bladder cancer (type of bladder cancer = "general")*

| Table 11. Model details for studies not focusing on a specific type of bladder cancer | | |
|---|---|---|
| **Author** | **Year** | **Model Details** |
| | | **Compartmental Models** |
| Baba, I. A. | 2019 | Models system of cells |
| Bois, F. Y., et al. | 1995 | Compartmental model; A physiological pharmacokinetic model was used to quantify the time course of the formation of the proximate carcinogen, N-hydroxy-4-ABP and the DNA-binding of the active species in the bladder. |
| Bunimovich-Mendrazitsky, S., et al. | 2016 | Compartmental model; They use a systems biology approach to describe the BCG-tumour-immune interplay and translate it into a set of mathematical differential equations. The variables of the equation set are the number of tumour cells, bacteria cells, immune cells, and cytokines participating in the tumour-immune response. |
| Farman, M., et al. | 2020 | Compartmental model; They developed the fractional-order immunotherapy bladder cancer model and used the BCG vaccine for treatment by using the Caputo fractional derivative operator ??(0,1]. A mathematical model has four variables B,E, Ti, Tu which represent the vaccine for the immune system, effector cells, total population of affected, and unaffected cells, respectively. |
| Guzev, E., et al. | 2019 | Compartmental model; Further optimize the mathematical model describing tumor–immune interactions in the bladder in response to combined therapy with BCG and IL-2, in light of newly collected data. In their model, 10 nonlinear ODEs govern the dynamics and the interactions among 10 different biological elements. |
| Ling, M. P., et al. | 2014 | Compartmental model; They present a probabilistic framework for assessing the human health risks from consuming raw and cooked fish that were cultured in groundwater Arsenic-contaminated ponds in Taiwan by linking a physiologically based pharmacokinetics model and a Weibull dose–response model. |
| Newton, M. A. | 1994 | Compartmental model; To determine which chromosome arms of bladder cancer cells (generated using an in vitro/in vivo transformation system) have an elevated loss probability (and thus putatively harbor suppressor genes), they construct a simple stochastic model of karyotype evolution. |
| Ooi, E., et al. | 2020 | Compartmental model, 3D model of mouse bladder; the study seeks to develop numerical models of mouse bladder to investigate the role of natural convection during bladder cancer treatment with GNR-assisted PTT (gold nanorods-assisted photothermal treatment). |
| Rechner, L.A., et al. | 2015 | Compartmental model; To calculate the excess relative risk for second cancer induction after radiotherapy for each tissue (ERRT) for each solution of beam weight combinations, they applied a risk model that accounts for initiation, inactivation, repopulation, and promotion (iirp), and the effects of fractionation. Additionally, in order to span the different possible relationships between dose and risk, they applied the linear-non-threshold risk model and 4 other non-linear risk models. |
| Rechner, L.A., et al. | 2012 | Compartmental model; Various risk models were combined with the dosimetric data (therapeutic and stray dose) to predict the excess relative risk (ERR) of cancer in the bladder and rectum. |
| schmit | 2018 | Compartmental model; The structural model (covariate-free model) consisted of a linear four-compartment PK (pharmacokinetic) model, with a zero-order input as infusion rate (previously published in [17]). The estimated PK parameters were: total body clearance (CL), volumes of distribution (V1, V2, V3 and V4) and three intercompartmental rate constants (K21,K31 and K41). |

| Table 11. Model details for studies not focusing on a specific type of bladder cancer | | |
|---|---|---|
| **Author** | **Year** | **Model Details** |
| Schneider, U. and B. Schafer | 2012 | Compartmental model; An existing model for cancer induction after fractionated radiotherapy which is based on cell mutations was extended here by including the effects of inflammation and proliferative stress, and an additional model parameter was established which describes acceleration. |
| Schooneveldt | 2020 | Experimental validation of a thermophysical fluid model on a cubic compartment constructed for the purpose |
| Suleiman, S.A., et al. | 2019 | Investigates the doses of radiation absorbed by different organs from cervical cancer treatment involving Co-60 source, and the related risk for cancer using computational phantoms (models used in computerized analysis) |
| Werneth, C. M., et al. | 2020 | Compartmental model; presents a general methodology for incorporating MCM (medical countermeasures) into the NASA Space Radiation Cancer Risk model and includes modifications of the background mortality rates (hazard rates) and the radiation risk coefficients to numerically quantify the benefits of MCM |
| Yoon | 2009 | A phantom (physical model) bladder was constructed and used to test the trajectory for optimal bladder imaging |
| Rodriguez-Brenes, I.A., et al. | 2017 | Compartmental model, human-derived bladder cancer xenografts in immunocompromised mice (cancer cells from humans implanted in mice); The central mathematical model tracks the following populations: Quiescent tumor stem cells, Q, actively proliferating tumor stem cells, S, transit amplifying tumor cells, T, differentiated tumor cells, D, and the concentration of wound-healing factors, W. This model therefore reflects the notion that a tumor is characterized by a basic cellular hierarchy that is similar to normal tissue. |
| Starkov, K.E. and S. Bunimovich-Mendrazitsky | 2016 | Compartmental model; dedicated to dynamical analysis of the tumor cell population model elaborated by Bunimovich-Mendrazitsky et al. in [3]. This model uses nine non-linear ODEs investigated numerically [3], in order to describe the effects of combined BCG and IL-2 immunotherapy for bladder cancer cells and upon other cells populations in order to evaluate comparative therapeutic scenarios. |
| Weiss, L. D., et al. | 2021 | Compartmental model; An ordinary differential equation model has been used to describe tissue hierarchy dynamics in a healthy tissue (Kunche et al., 2016; Lander et al., 2009), and the models presented here build on these approaches. While cell lineages consist of stem cells, transit amplifying cells, and terminally differentiated cells, our models make a simplification and take into account only stem cells (which encompass all the proliferating cells) and differentiated cells. It has been shown that introduction of negative feedback loops can result in more realistic behavior, where a stable equilibrium is attained for P>0.5 (Lander et al., 2009). This was shown in the context of two specific feedback loops, and subsequently generalized to comprehensively list all possible (positive and negative) feedback loops compatible with stability (Komarova, 2013; Komarova and van den Driessche, 2017; Yang et al., 2017). Here, we also use a general model to assume different kinds of feedback on the rate of cell division, L, the rate of cell death, D, and the probability of self-renewal, P. We also add the possibility that stem cells die with a rate ?(which can also be subject to feedback). In the context of our model, feedback is equivalent to a dependence of rates and probabilities on the population sizes, x (stem cells) and/or y (differentiated cells). |

| Table 11. Model details for studies not focusing on a specific type of bladder cancer | | |
|---|---|---|
| **Author** | **Year** | **Model Details** |
| **Cohort-based simulation models** | | |
| Geller, A. C., et al. | 1990 | Evaluates a screening program model |
| Neafsey, P., et al. | 2009 | The study is focused on the allele frequencies of polymorphysims of a cytochrome that have been suspected to be associated with increased bladder cancer risk in different populations. Agressive (stage III) and non-aggressive cancer both mentioned, but whether MIBC and/or NMIBC not mentioned. |
| Schorp, M.K. and D.E. Leyden | 2010 | Study is focused on determining the impact of environmental tobacco smoke on a known bladder carcinogen (4-ABP). No specific type of bladder cancer mentioned. |
| Walker, K., et al. | 2009 | The study is focused on the allele frequencies of polymorphysims of two enzymes that have been suspected to be associated with increased bladder cancer risk in different populations. No specific type of bladder cancer mentioned. |
| **Decision tree models** | | |
| Corwin, H. L. and M. D. Silverstein | 1988 | Aims to determine performance of different strategies in the diagnosis of neoplasia (no specific bladder cancer type) |
| Halpern, J. A., et al. | 2017 | Evaluates the cost-effectiveness of 4 different diagnostic methods used to investigate asymptomatic microscopic hematuria |
| J. Bandari, M. E. Nielsen, B. L. Jacobs and K. J. Smith | 2018 | Evaluates the cost-effectiveness of an evaluation of asymptomatic microhematuria |
| Martin, A. D., et al. | 2011 | Study focused on a cost analysis comparing robot-assisted radical cystectomy and open radical cystectomy. Type of bladder cancer was not of interest/not metioned |
| Svatek, R. S., et al. | 2006 | Focuses on the evaluation of the cost associated with screening for bladder cancer using a non-invasive tumor marker |
| **Microsimulation models** | | |
| Cheong, J. K. K., et al. | 2021 | In silico model, mouse bladder |
| Dietsch, A., et al. | 2000 | In vivo study on anesthetized dogs |
| Ellwein, L. B. and S. M. Cohen | 1988 | Models system of cells, rat |
| Georgieva, M., et al. | 2019 | Assesses 5 different guidelines for the initial evaluation of hematuria |
| Greenfield, R. E., et al. | 1984 | Models system of cells |
| Grieco, L., et al. | 2013 | Creates a logical model corresponding to the MAPK network and its influence on cancer cells |
| Krishnan, N., et al. | 2016 | Study focused on individuals who undewent radical cystectomy for baldder cancer. Type of bladder cancer was not of interest/mentioned. |

### S.8 Technical details about simulation models

| Table 12. Number of studies (and % over the total N=97 eligible studies) providing technical details on each methodology used for the development of the simulation models they use. | |
|---|---|
| **Technical details included** | **n (%)** |
| Calibration | 9 (9.3) |
| Validation | 32 (32.9) |
| Sensitivity Analysis | 40 (41.2) |
| State Diagram | 55 (56.7) |
| Code available | 4 (4.12) |

### S.9 Statistical Software used for simulation models

| Table 13. Statistical software used for the development of simulation models for bladder cancer. | |
|---|---|
| **Software** | **N** |
| Not mentioned | 38 |
| TreeAge | 17 |
| MATLAB | 11 |
| R | 3 |
| C++ | 2 |
| Crystal Ball | 2 |
| Microsoft Excel | 2 |
| ARCHER MC code, MCNPX MC code | 1 |
| AngularJS, Microsoft Azure, R | 1 |
| Archimedes Model | 1 |
| COMSOL Multiphysics | 1 |
| CellDesigner, GINsim | 1 |
| CellNetAnalyzer, ImageJ | 1 |
| Compucell3D | 1 |
| Eclipse (Varian Medical Systems, Palo Alto, CA, USA), MATLAB | 1 |
| GINsim, R | 1 |
| MATLAB, ImageJ, SolidWorks, COMSOL Multiphysics | 1 |
| MATLAB, Microsoft Excel | 1 |
| MATLAB, PCIBIRD CUBES | 1 |
| Microsoft Excel | 2 |
| NONMEM, SAS | 1 |
| Oracle | 1 |
| R, TreeAge | 1 |
| S | 1 |
| SAS | 1 |
| SAS, TreeAge | 1 |
| Sigma-HyperPlan | 1 |
| Simul8, Stat-Fit | 1 |
| StatXact | 1 |
| **Total** | **97** |

### S.10 Objectives and Conclusions of simulation studies

Table 14. Objectives and conclusions of eligible simulation studies for bladder cancer.

| Year | Author | Model Type | Objective | Conclusion |
|---|---|---|---|---|
| 2017 | Aboulaich, R., et al. | Compartmental model | Find optimal BCG dose for minimizing the total number of tumors in the presence of diffusion process for superficial bladder cancer. | The study suggests an optimal control approach for fast eradication of tumor cells and demonstrates a promising potential of BCG bacterium as an effective immunotherapeutic agent even in the presence of biological stochasticity. |
| 2015 | Al Awamlh, B. A. H., et al. | Decision tree | CEA comparing different treatments (fulguration vs TURBT) in terms of ICER and QUALYs | Office-based cystoscopy and filguration was more cost-effective compared to TURB for low-risk NMIBC |
| 2006 | Avritscher, E. B., et al. | Cohort-based | Estimate the lifetime cost of bladder cancer and the contribution of complications to the total costs, and suggest more cost-effective surveillance strategies and approaches for preventing complications to reduce BCA clinical and economic consequences. | The study provides evidence of the need of more cost-effective surveillance strategies and approaches for preventing complications to minimize the clinical and economic consequences of the disease. |
| 2019 | Baba, I. A. | Compartmental model | Evaluate the effectiveness of BCG in controlling superficial BCA based on disease-free and endemic equilibrium points. | Results from numerical simulations indicated that the greater the tumor growth rate the faster the tumor spreads and is present in cells. |
| 2018 | Bandari, J., et al. | Decision tree | Cost-effectiveness analysis to compare upfront vs confirmed evaluation (requiring confirmed UA before initiating full evaluation). | The study provides a compelling argument in favor of sex stratification and confirmatory urinalysis in specific circumstances. |
| 1995 | Bois, F. Y., et al. | Compartmental model | Investigate the impact of interindividual heterogeneity in the metabolism of 4-aminobiphenyl (ABP) and in physiological factors on human cancer risk | The study showed that the four factors contributing most significantly to interindividual differences in DNA binding of ABP in human bladder are urine pH, ABP N-oxidation, AE3P N-acetylation and urination frequency. |
| 2007 | Bunimovich-Mendrazitsky, Svetlana, Eliezer Shochat, et al. | Compartmental model | Understand the details of the process and impact of BCG on treating superficial BCA based on multiple equilibrium points. | The study showed that treatment intensity must be kept in limited bounds as, while small levels may fail to clear the tumor, large can lead to an over-stimulated immune system with dangerous side effects for the patient. |
| 2016 | Bunimovich-Mendrazitsky, Svetlana, et al. | Compartmental model | Explore the effect of time-honored protocols for treating NMIBC with BCG installations and combinations with IL-2 to determine a more effective personalized protocol, taking into account tumor size (volume) and tumor growth (grade). | Model validated to clinical trial results. The results showed that subpopulation of non-responsive patients may benefit from an intensified combined BCG and IL-2 maintenance treatment for preventing tumor recurrence. |
| 2008 | Bunimovich-Mendrazitsky, Svetlana, Helen Byrne, et al. | Compartmental model | Explore the effectiveness of BCG and determine threshold values of BCG instillation dose and rate for successfully treating superficial BCA. Also identify treatment regimes resulting in tumor destruction, at low levels of undesirable side effects maintained by the immune system. | The study demonstrates the effectiveness of BCG treatment, the response of which depends on the initial tumor size and state. Extensions of the model can form the basis for future mathematical studies that will inform clinical practice. |
| 2011 | Bunimovich-Mendrazitsky, Svetlana, Jean Claude Gluckman, et al. | Compartmental model | Explore the effectiveness of BCG, alone or combined with IL-2 after surgical removal of tumors, evaluating dynamical outcomes from interactions between immune and tumor cells, to determine conditions that would result in successful BCA treatment. | Combination of BCG with IL-2 trt will not necessarily result in better results for treating BCA patients |

| Table 14. Objectives and conclusions of eligible simulation studies for bladder cancer. | | | | |
|---|---|---|---|---|
| Year | Author | Model Type | Objective | Conclusion |
| 2015 | Bunimovich-Mendrazitsky, Svetlana, Vladimir Pisarev, et al. | Compartmental model | A model to describe the initiation and progression of a low-grade urinary bladder carcinoma within a framework of urothelial cell dynamics with emphasis on the 2 common types of BCA; bladder polyps and carcinoma in situ. | Analysis of histological structure of bladder tumor is important to avoid misdiagnosis and wrong treatment. The model is a valuable tool for describing BCA progression due to exposure to carcinogens and the oxygen dependent expression of genes promoting tumor growth. |
| 2011 | Bunimovich-Mendrazitsky, Svetlana, Yakov Goltser, et al. | Compartmental model | Explored the immune response in BC as an effect of BCG treatment. | Results from this work can be used for effective and efficient design of relevant clinical protocols. |
| 2019 | Chalk, D., et al. | Microsimulation | Identify key bottle necks in the bladder cancer pathway and evaluate healthcare alternatives for a more effective diagnosis and treatment for non-metastatic MIBC. | The key bottlenecks identified in this study were 2 waits for a patient; 1) to receive TURBT and 2) to be contacted by a nurse specialist to discuss diagnosis. The model simulates the situation at the Royal Cornwall Hospital. |
| 2021 | Cheong, J. K. K., et al. | Microsimulation | Investigate the efficacy of GNR-PTT in the treatment of bladder cancer in mice for tumours growing at three different locations on the bladder; closest to skin surface (Case 1), bottom half of the bladder (Case 2), and side of the bladder (Case 3), based on an in-silico approach. | Results showed that the tumour's location can significantly affect GNR_PPT treatment efficacy |
| 1988 | Corwin, H. L. and M. D. Silverstein | Decision tree | Evaluate the performance of alternative strategies involving sequences of diagnostic tests for the diagnosis of neoplasia in adults with asymptomatic microscopic hematuria, including ultrasound, excretory, urography, angiography, computerized tomography and cystoscopy. | Strategies using cystoscopy or ultrasound as the initial diagnostic test minimized cost and morbidity while maintaining diagnostic accuracy. Excretory urography does not add significantly to diagnostic accuracy, but it does add to cost and morbidity. |
| 2019 | Criss, S. D., et al. | Microsimulation | Evaluate the effect of PD-L1 testing on the cost-effectiveness of pembrolizumab for second-line treatment (after first-line platinum-based chemotherapy) of advanced urothelial carcinoma in the bladder from the U.S. societal perspective. The study compares 3 strategies; 1) all treated with pembrolizumab, 2) all treated with chemotherapy, 3) use PD-L1 test results to determine if the patient should be treated with pembrolizumab or chemotherapy | The study showed that pembrolizumab was not a cost-effective option for the treatment of advanced urothelial carcinoma of the bladder for either of the strategies tested at a WTP threshold of $100,000/QALY. PD-L1 testing to select patients who may have better associated outcomes may improve the affordability of pembrolizumab. |
| 2016 | Dansk, V., et al. | Cohort-based | CEA to compare HAL BLFC as adjunct to WLC for detection and treatment of NMIBC compared to WLC alone. | HAL BLFC allowed more outpatient treatment with improved recurrence detection and reduced transurethral resection of the bladder tumors, cystectomies, bed days and operating room time, with minimal cost impact across all risk groups, demonstrating the economic benefits of introducing HAL. |
| 2000 | Dietsch, A., et al. | Other | Evaluate the performance of microwave thermotherapy and determine the therapeutic protocol associating chemotherapy and thermotherapy in the treatment of the bladder cancer (vesical carcinoma). | Results from this study suggest the clinicians to determine a new operative record for the treatment of bladder cancer using an association between chemotherapy and thermotherapy |
| 1988 | Ellwein, L. B., and S. M. Cohen | Microsimulation | Simulate the carcinogenesis process through cellular kinetics and changes in the urinary bladder, as well as the association of tumor incidence with the timing and magnitude of changes to cellular kinetics. | It is demonstrated that response in rats following administration of nongenotoxic compounds, such as sodium saccharin, can be explained entirely on the basis of cytotoxicity and consequent hyperplasia alone. |
| 1984 | Ellwein, L. B., et al. | Microsimulation | MSM to evaluate screening protocols for bladder cancer with applications to CEA | The study suggests that use of cytologic studies in a screening program in an appropriately selected asymptomatic population would |

| Table 14. Objectives and conclusions of eligible simulation studies for bladder cancer. | | | | |
|---|---|---|---|---|
| Year | Author | Model Type | Objective | Conclusion |
| | | | | reduce the number of bladder cancer deaths and extend life for those discovered to have the disease. Recommendations for inaugurating a widespread screening program, require more extensive CEAs to determine whether the anticipated gains from such a program justify incurred costs. |
| 1988 | Ellwein, Leon B., and George M. Farrow | Decision tree | Evaluate the potential of voided-urine cytology as a screening test for the early detection of urinary bladder cancer in asymptomatic populations. | Analyses predict that the predisposition of cytology screening to identify the high-grade, aggressive form of the disease will result in gains in life expectancy, and except for the risk of a false-positive outcome, it compares favorably with what could theoretically be obtained if a 100% accurate screening test were available. |
| 2020 | Farman, M., et al. | Compartmental model | A biological model to evaluate the effect of BCG vaccine for BCA treatment. | The study showed promising results of the BCG vaccine for controlling the disease at the initial stage to overcome the risk of living with cancer. |
| 2017 | Galsky, M. D., et al. | Microsimulation | Evaluate the performance of NAC for treating MIBC using a web-based tool for shared decision making, in an effort to address particular challenges of decision making, including contextualization of the potential benefits and communication of this information in an understandable manner. Current efforts are focused on acceptability testing among patients and physicians in the clinical setting and ultimately testing to establish clinical utility of an SDM protocol for patients with MIBC before cystectomy. | The NAC therapy showed the standard of care for communicating the potential benefit of NAC for MIBC is undefined |
| 1990 | Geller, A. C., et al. | Cohort-based | Evaluate performance of 4 screening tests (i. Voided Urine Cytology, ii. QFIA, iii. Routine Urinalysis for Microhematuria, iv) Home Dipstick Testing) and define a protocol to maximize case detection and minimize the number of invasive procedures by utilizing both flexible and rigid cystoscopy. | Transitional cell carcinoma of the bladder appears to have a latent, preclinical phase during which screening for its detection can be effective. Detecting preinvasive disease at this point could decrease the proportion of cases associated with superficial bladder cancer that recur or progress to advanced disease. The study suggests that simultaneous tests can increase the ability to detect lesions with invasive potential at a preclinical stage. |
| 2019 | Georgieva, M., et al. | Microsimulation | Compare advantages, harms, and costs associated with 5 guidelines for hematuria evaluation; i) Dutch, ii) Canadian Urological Association (CUA), iii) Kaiser Permanente (KP), iv)Hematuria Risk Index (HRI), and v)American Urological Association (AUA). | The study indicates that uniform CT imaging for patients with hematuria was associated with increased costs and harms of secondary cancers, procedural complications, and false positives, with only a marginal increase in cancer detection. Risk stratification may optimize the balance of advantages, harms, and costs of CT. The study suggests that, in addition to its substantial costs, the potential harms of the intensive application of uniform CT urography may outweigh the advantages of early diagnosis of urinary tract malignant neoplasms. |
| 2006 | Grabnar, I., et al. | Compartmental model | Understand and describe the bladder pharmacokinetics to optimize intravesical drug delivery in the tumor and its vicinity and reduce systemic toxicity, for treating superficial BCA. | Both chitosan (CH) and polycarbophil (PC) polymers increase permeability of the bladder wall by diffusion enhancement in the urothelium and presumably by improving the contact with the bladder surface. |

| Table 14. Objectives and conclusions of eligible simulation studies for bladder cancer. | | | | |
|---|---|---|---|---|
| Year | Author | Model Type | Objective | Conclusion |
| 2013 | Green, D. A., et al. | Cohort-based | Compare cost-effectiveness of fulguration vs transurethral resection of bladder tumour (TURBT) with and without perioperative intravesical chemotherapy (PIC) for managing low-risk carcinoma not invading bladder muscle (NMIBC). | Fulguration without PIC was the most cost-effective strategy for treating low-risk NMIBC. The effectiveness of PIC and the cost of TURBT can materially impact the cost effectiveness of the different management strategies. |
| 1984 | Greenfield, R. E., et al. | Microsimulation | Mathematical model to simulate the carcinogenesis process in two stages and explain and describe the impact of exposure to nongenotoxic compounds (sodium saccharin) and FANFT on this process. | FANFT seems to have a direct effect (increase) on the probability of cell initiation, cell population and mitotic rates, unlike sodium saccharin which seems to only affect the stem cell population and mitotic rates. |
| 2013 | Grieco, L., et al. | Microsimulation | Develop a biological model to describe the impact of environmental conditions on the carcinogenesis process. | Results consistent with published data. In silico experiments further delineate the roles of specific components, cross-talks and regulatory feedback in cell fate decision, and suggest that established bladder cancer deregulations (e.g., Epidermal Growth Factor Receptor (EGFR) over- expression and Fibroblast Growth Factor Receptor 3 (FGFR3) activating mutations) can be connected proliferative or anti-proliferative mechanisms. |
| 2019 | Guzev, E., et al. | Compartmental model | Compare treatment protocols (BCG installation vs BCG combined with IL-2) in the presence of superficial bladder cancer. | A program that can be used as an auxiliary tool for doctors in the decision-making process of choosing individual treatment for a BC patient. |
| 2017 | Halpern, J. A., et al. | Decision tree | Cost effectiveness analysis to compare 4 common diagnostic approaches for evaluating AMH; (1) computed tomography (CT) alone; (2) cystoscopy alone; (3) CT and cystoscopy combined; and (4) renal ultrasound and cystoscopy combined. | The combination of renal ultrasound and cystoscopy is the most cost-effective among 4 diagnostic approaches for the initial evaluation of AMH. The use of ultrasound in lieu of CT as the first-line diagnostic strategy will optimize cancer detection and reduce costs associated with evaluation of AMH. The findings indicate a need to critically evaluate the appropriateness of current clinical practices and potentially change in guidelines to reflect the most effective screening strategies for patients with AMH. |
| 2019 | Heijnsdijk, E. A. M., et al. | Microsimulation | Cost-effectiveness analysis to compare surveillance schedules (16 in total - all combinations of 3, 6, 12, or 24 months for 2, 5, and 10 years) for non-muscle-invasive bladder cancer (NMIBC) amongst older adults (65-85 years old). | Cystoscopy-based surveillance for NMIBC is cost-effective for younger patients (<75 years old) suggesting need for age-specific surveillance recommendations. |
| 2021 | Hird A., et al. | Microsimulation | Compare 3 treatment pathways in UTUC: nephroureterectomy (NU) alone, neoadjuvant chemotherapy (NAC), and adjuvant chemotherapy (AC) using a microsimulation model. | The study supports increased use of NAC in UTUC until robust randomized trials are completed. The ultimate choice should be based on patient and tumor factors. |
| 2017 | Kanigel Winner, K. R. and J. C. Costello | Microsimulation | Simulate the biological process of lung cancer metastasis from a primary bladder tumor due to resistance of cancer cells to chemotherapy. | NA |
| 2013 | Kashdan, E. and S. Bunimovich-Mendrazitsky | Compartmental model | Simulate the biological mechanism (carcinogenesis) of tumor invasion and examine pathways leading to invasive BC. Carcinogenesis starts when the chemicals known as carcinogens penetrating from the bladder lumen affect top(umbrella) cells of the urothelium. | NA |
| 1989 | Kent, D. L., et al. | Microsimulation | Exploring optimal scheduling of cystoscopies to improve efficiency in monitoring patients for recurrent superficial bladder cancer. | Results suggest that a formal optimization approach to scheduling, considering histologic grade, tumor stage, and number of prior |

| Table 14. Objectives and conclusions of eligible simulation studies for bladder cancer. | | | | |
|---|---|---|---|---|
| Year | Author | Model Type | Objective | Conclusion |
| | | | | tumors as predictive factors, can improve the efficiency of monitoring patients for recurrent superficial bladder cancer. |
| 2021 | Khaki, A. R., et al. | Decision tree | Cost-effectiveness to compare ICI with CBC therapies for MIBC. Primary analysis: pembrolizumab vs ddMVAC. Secondary analysis: atezolizumab or nivolumab/ipilimumab vs gemcitabine/cisplatin. | ICIs were not cost-effective as neoadjuvant therapies, except when atezolizumab was compared with ddMVAC. Randomized clinical trials, larger sample sizes and longer follow up are required to better understand the value of ICIs as neoadjuvant treatments. |
| 2017 | Khan, F. M., et al. | Compartmental model | Simulate the carcinogenesis process using the regulatory network around E2F, a family of transcription factors whose deregulation has been associated to cancer progression, chemoresistance, invasiveness, and metastasis. | The integrative network-based methodology, exemplified in the case of E2F1-induced aggressive tumors, has the potential to support the design of cohort- and tumor type-specific treatments to fight metastasis and therapy resistance. |
| 2011 | Kokorowski, P. J., et al. | Microsimulation | Cost-effectiveness analysis to compare usual care (annual physician visit with an ultrasound examination of the kidneys and augmented bladder) with the same care as well as annual cystoscopic and cytological evaluation of the augmented bladder for patients with spina bifida. | Annual screening for malignancy among patients with spina bifida with cystoplasty using cystoscopy and cytology is unlikely to be cost effective at commonly accepted willingness to pay thresholds. Conclusion sensitive to a higher than expected risk of malignancy and highly optimistic estimates of screening effectiveness. |
| 2019 | Kongnakorn, T., et al. | Cohort-based | Cost analysis of avelumab as a second-line (2L) treatment option for patients with locally advanced or mUC from the perspective of a US third-party payer (commercial and Medicare). | Study results demonstrate cost-neutral budget impact of avelumab (as a 2nd line trt) on patients with locally advanced or mUC, for both commercial and Medicare health plans in the US. |
| 2016 | Krishnan, N., et al. | Microsimulation | Determine optimum outpatient follow up regimens (combinations of office visits & phone calls, and timing (days) after discharge) for radical cystectomy. | Timing and number of outpatient encounters are key determinants in improving effectiveness of outpatient follow up care for patients treated with radical cystectomy and can help with reducing readmission burden in the population. |
| 2020 | Kukreja, J. B., et al. | Decision tree | Cost-effectiveness analysis to compare RARC with open cystectomy (OC) for bladder cancer. | RARC is cost-effective compared to OC when the rates of complications and transfusions are significantly lower. |
| 2007 | Kulkarni, GS , et al | Microsimulation | Decision analysis to compare 2 treatments for high-risk superficial (stage T1; grade G3) bladder cancer in terms of LE and QALE; (1) immediate nerve-sparing cystectomy with orthotopic neobladder creation vs (2) conservative therapy with potential for delayed cystectomy. | Results demonstrated that younger patients with high-risk T1G3 bladder had a higher LE and QALE with immediate cystectomy. |
| 2021 | Lazebnik, T., et al. | Compartmental model | Describe the BCG immunotherapy dynamic taking into consideration an approximation of the bladder's geometry using PDE. | The proposed model takes into account the initial distribution of the cancer cells in the geometry of the bladder and as such can provide more customized treatment by providing tumor polyp depth in the urothelium. In addition, it can be used to explore time optimal treatment protocol for the average case and recover-rate optimal, personalized treatment protocol based on initial tumor distribution. |
| 2012 | Lee, C. T., et al. | Decision tree | CEA to estimate the economic and humanistic gains associated with preventable recurrences of TURBT combined with immediate IVIC for treating patients initially diagnosed with NMIBC. | Use of immediate intravesical chemotherapy in the US has the potential to substantially decrease the economic and humanistic burdens of NMIBC. |
| 2014 | Ling, M. P., et al. | Compartmental model | Evaluate the human health risks from consuming raw and cooked fish cultured in groundwater As-contaminated ponds in Taiwan combining a PBPK with a Weibull dose-response model. | Cooking increases the As concentration in contaminated fish, but baking resulted in lower ('acceptable') risks and is recommended as cooking method, unlike frying which associated with significant (beyond acceptable) health risks. |

| Table 14. Objectives and conclusions of eligible simulation studies for bladder cancer. | | | | |
|---|---|---|---|---|
| Year | Author | Model Type | Objective | Conclusion |
| 2002 | Lotan, Yair, and Claus G. Roehrborn | Decision tree | CEA to compare follow up protocols for superficial TCC after TURBT | A modified follow up protocol in TCC cases using a urine based tumor marker alternating with cystoscopy and/or cytology can be more cost-effective than the standard care. |
| 2006 | Lotan, Yair, et al. | Cohort-based | CEA to evaluate screening of high-risk patients for bladder cancer using urine-based markers. | Urine-based markers seem to be cost-effective in a high-risk population. Prospective RCTs in completely asymptomatic high-risk cohorts are indicated before bladder cancer screening can be recommended. |
| 2020 | Magee, D., et al. | Microsimulation | Compare radical cystectomy (the historic gold-standard treatment for MIBC) with trimodal therapy in terms of QALE, LE, and BC recurrence. | RC results in a longer LE compared to TMT (0.54 years), but with a lower QALE (-0.07 years). The preferred treatment strategy varied with patient age. |
| 2009 | Malmstrom, P. U., et al. | Cohort-based | Estimate the budget impact on the Swedish health service of using HAL in conjunction with WLC in the management of NMIBC for 1 year following initial diagnosis. | HAL cystoscopy (as an adjunct to white light in guiding TURB) may result in reduction of invasive, time-intensive and high-cost procedures such as cystectomy and TURB, compared with WLC alone for treating NMIBC. |
| 2000 | Marchetti, A., et al. | Decision tree | Estimate 1st- and 2nd-year clinical costs associated with intravesical valrubicin therapy for patients with Bacilli Calmette- Guerin (BCG) - refractory carcinoma in situ (CIS) of the urinary bladder. | Cost-consequence analysis indicates valrubicin therapy as a viable alternative to the standard RC for patients with CIS who have poor prognosis (due to advanced age or comorbidities) or prefer medical rather than surgical management. |
| 2011 | Martin, A. D., et al. | Decision tree | Cost analysis comparing robot assisted radical cystectomy (RARC) versus open radical cystectomy (ORC) based on single-institution data. | RARC can be a cost-effective alternative due to higher complication rates of ORC, with operative time and length of stay being the most critical cost determinants. |
| 2018 | Alkama, et al. (2018) | Compartmental model | Explore optimal control approach based on optimal BCG dosage and duration of treatment for superficial bladder tumors. | NA |
| 2019 | Michels, C. T. J., et al. | Decision tree | Cost-effectiveness, comparing RARC to ORC for MIBC | Although RARC is more expensive than ORC it may result in fewer complications. |
| 2000 | Nam, R. K., et al. | Decision tree | Cost analysis to compare cystoscopy and cytology (standard care) vs urinary markers (modified care) for long-term surveillance of NMIBC. | Urinary marker testing (from the payer-perspective) is less expensive for (3 year) follow up of patients with NMIBC compared to the standard method of cystoscopy and urinary cytology, with the difference increasing with surveillance time. |
| 2019 | Nave, O., et al. | Compartmental model | SPVF model to evaluate a combination of BCG and IL-2 for bladder cancer treatment. | The model provides explicit expressions of equilibrium points and allows investigation of the stability of these points. |
| 2009 | Neafsey, P., et al. | Cohort-based | Analysis to characterize (a) influence of genotype on phenotype based upon in vivo metabolism studies of probe drugs and (b) frequency of the major genotypes in different population groups. Improve understanding of the variability in CYP2D6 function possible in the population for assisting quantitative risk assessments (using PBPK modeling) and related variability in internal dose for substrates whose activation or detoxification is mediated by CYP2D6. | CYP2D6 is the primary enzyme responsible for the metabolic activation or detoxification of a particular xenobiotic. Focus on CYP2D6 in metabolism screens of environmental toxicants would help risk assessors determine the role polymorphisms may play in modulating internal dose and toxic response. |
| 1994 | Newton, M. A. | Compartmental model | Simulation model to describe chromosome loss during tumour progression and predict possible locations of bladder cancer suppressor genes. | The model indicates significant losses on chromosomes 3, 18, 13, 10, 11, and y related to bladder cancer progression. |

| Table 14. Objectives and conclusions of eligible simulation studies for bladder cancer. | | | | |
|---|---|---|---|---|
| Year | Author | Model Type | Objective | Conclusion |
| 2014 | Newton, P. K., et al. | Cohort-based | Simulate the dynamics of metastatic cancer and demonstrate how entropy and graph conductance can be used in the context of metastatic spread to quantify, compare and co-group the most prevalent cancer types worldwide. | The models suggest that grouping cancers according to their entropy values can provide a useful framework for characterizing and simulate metastatic cancer in terms of predictability, complexity and metastatic potential. |
| 2020 | Ooi, E., et al. | Compartmental model | Investigate the effects of natural convection inside the bladder and at skin surface during GNR-PTT therapy based on examination of bladder cancer in mice. | NA |
| 2020 | Parmar, A., et al. | Decision tree | Cost-utility analysis (CUA) to compare atezolizumab with chemotherapy as second-line treatment for metastatic bladder cancer. | Atezolizumab is not considered cost-effective for the second line treatment of mBC. |
| 2015 | Patel, S., et al. | Microsimulation | Application of the Archimedes computational model to compare RC versus intravesical MMC therapy with respect to clinical and economic outcomes for BCG-refractory NMIBC (<cT2 disease). | The simulations did not reveal a dominant trt. Results showed overall that RC is more cost-effective than MMC. Archimedes and similar simulation models are proposed as helpful tools for feasibility assessment of dedicated RCTs . |
| 2012 | Porta, N., et al. | Microsimulation | Simulation model to predict the risk of progression for patients diagnosed with BCA. | The proposed simulation model can be a helpful tool for subject-specific management of patients with BCA. |
| 2015 | Rechner, Laura A., John G. Eley, et al. | Compartmental model | Demonstrate feasibility of risk-optimized proton therapy and determine the combination of beam angles and fluence weights that minimize the risk of second cancer in the bladder and rectum for a prostate cancer patient. | NA |
| 2012 | Rechner, Laura A., Rebecca M. Howell, et al. | Compartmental model | Compare the predicted risk of cancer following proton arc therapy and VMAT for prostate cancer, and predict the ERR of cancer in the bladder and rectum. | Proton arc therapy significantly reduces the predicted risk of radiogenic second cancer in the bladder and rectum following prostate radiotherapy compared with that following VMAT. |
| 2015 | Remy. E., et al. | Microsimulation | Simulation modeling to describe patterns of genetic alterations in patients with bladder tumors (MIBC or NMIBC). | Work helpful for predicting combinations of major gene alterations leading to invasiveness through two main progression pathways in bladder cancer influence. |
| 2013 | Rentsch, C. A., et al. | Microsimulation | Evaluate optimal approaches for treating NMIBC with BCG given 4 main clinical factors : (1) duration between resection and the first instillation; (2) dose; (3) indwelling time; and (4) treatment interval of induction therapy, using cure rate as the primary endpoint. | Findings suggest that a rigorous time management and instillation plan with extended treatment intervals can be crucial for successful BCG treatment of NMIBC for patients at high risk of recurrence. |
| 2017 | Rodriguez-Brenes, I.A., et al. | Compartmental model | Identify determinants of BCG treatment response for superficial BCA in patients with high risk of recurrence, accounting for interactions between BCG, the immune system, the bladder mucosa and tumor cells. | Extending the duration between the resection and the first BCG instillation negatively influences treatment outcome. Higher BCG doses and longer indwelling times may improve the probability of tumor extinction. An inter-instillation interval two times longer than the seven-day interval used in the current standard of care would substantially improve treatment outcome. |
| 2019 | Royce T., et al. | Microsimulation | CEA to compare TMT vs RC for treating MIBC for patients 67 years old, diagnosed at clinical stage T2-T4aN0M0. | Treatment of MIBC with organ-sparing TMT can be cost-effective for appropriately selected patients (gain of QALYs) relative to RC. |
| 2014 | Rubio, G., et al. | Microsimulation | Flowgraph model to simulate the evolution of NMIBC. | A flowgraph model is a helpful, flexible tool for describing NMIBC evolution considering multiple recurrences and important covariates. |
| 2018 | Saad, Farouk Tijjani, and Evren Hincal | Compartmental model | Study the dynamics and identify optimal BCG dose required for treating superficial BC, to minimize the cancer cells and action of the immune | NA |

| Table 14. Objectives and conclusions of eligible simulation studies for bladder cancer. | | | | |
|---|---|---|---|---|
| Year | Author | Model Type | Objective | Conclusion |
| | | | checkpoints, as well as maximizing the number of immune and normal cells in the bladder. | |
| 2017 | Saad, Farouk Tijjani, et al. | Compartmental model | Study the dynamics of immune suppressors/checkpoints, immune system, and BCG in the treatment of superficial bladder cancer. | NA |
| 2016 | Sadee, C. and E. Kashdan | Compartmental model | Mathematical model to simulate the physical process of the effect of chemo-thermotherapy on treating NMIBC cases with high risk of recurrence. | Promising results of C-HT for treating NMIBC also given new types of thermal delivery systems such as the SB-TS 101 device. |
| 2009 | Santamaria, C., et al. | Microsimulation | A Markovian approach to simulate superficial vesical carcinoma progression taking into account up to two recurrences. | The applied methodology allows to distinguish between different types of events: recurrence and progression. Results showed that grade is highly significant for progression. |
| 2018 | Sarfaty, M., et al. | Decision tree | CEA comparing pembrolizumab with chemotherapy for treating second-line advanced bladder cancer from the perspective of payers in multiple countries (US, UK, Canada, and Australia). | With standard WTP thresholds, pembrolizumab may be considered cost-effective in the US (due to high US drug prices leading to the highest ICER) but not in the other countries examined. |
| 2018 | Schmitt, Antonin, et al. | Compartmental model | PK and PD analysis of Vinflunine, a novel tubulin-targeted inhibitor indicated as a single agent for the treatment of bladder cancers after failure of prior platinum-based therapy. | Results from this global comprehensive clinical pharmacological analysis for intravenous vinflunine can help with drive dose adjustment. |
| 2012 | Schneider, U. and B. Schafer | Compartmental model | In the present study, an existing model for cancer induction after fractionated radiotherapy which is based on cell mutations was extended by including the effects of inflammation and proliferative s | It is proposed that tissue injury due to high doses of radiation may be due to enhanced cell proliferation. Additional research is needed to understand the impact of accelerated carcinogenesis for radiotherapy patients. |
| 2020 | Schooneveldt, G., et al. | Compartmental model | Assess the accuracy of a thermophysical fluid model for hyperthermia treatment planning near fluid volumes in the body. | Sufficient accuracy of the model, indicating that it can be helpful for evaluating treatment planning and monitoring of BCA. |
| 2016 | Schooneveldt, G., et al. | Compartmental model | Improved mathematical dielectric and thermophysical model of the urinary bladder to evaluate the effectiveness of hyperthermia treatment planning. | Modeling convection in the urinary bladder is a key factor for accurate hyperthermia treatment planning in the pelvic area. |
| 2010 | Schorp, M.K. and D.E. Leyden | Cohort-based | A combination of toxicokinetics, exposure modeling, and Monte Carlo simulation to evaluate the impact 4-ABP-Hb adduct levels from exposure to ETS on the risk of bladder cancer. | NA |
| 2020 | Sharma, V., et al. | Decision tree | CEA of BCG maintenance for patients with intermediate and high risk NMIBC, in view of BCG shortage. | BCG not cost effective for maintenance of the entire population. Priority should be high-risk NMIBC cases. Findings in accordance to the AUA guidelines to allocate BCG for induction rather than maintenance. |
| 2016 | Starkov, K.E. and S. Bunimovich-Mendrazitsky | Compartmental model | Validation of the Bunimovich-Mendrazitsky (2015) mathematical model for studying the immune system response to combined therapy for bladder cancer with BCG and IL-2. | Model validation of a nine-dimensional BCA immunotherapy, also obtaining global tumor clearance conditions via the localization method of compact invariant sets. |
| 2019 | Suleiman, S.A., et al. | Compartmental model | The present work aimed to evaluate organ doses and related risk for cancer from external beam radiation treatment (EBRT) and high-dose-rate (HDR) brachytherapy (BT) involving Co-60 source for patients with cervical carcinoma in Tanzania based on Monte Carlo methods and to evaluate the secondary cancer risks in their lifetime. | The chances of developing secondary cancer take years following radiation therapy are extremely low, but the results of present study can support to establish a future database on secondary cancer risks involving radiation therapy in patients with cervical cancer by using HDR-BR and EBRT with Co-60 source in Tanzania and other developing countries. |

| Table 14. Objectives and conclusions of eligible simulation studies for bladder cancer. | | | | |
|---|---|---|---|---|
| Year | Author | Model Type | Objective | Conclusion |
| 2018 | Sutton, A. J., et al. | Decision tree & Microsimulation | This study examines whether an alternative new urine-based diagnostic test, the DCRSHP, is cost-effective as a triage diagnostic tool compared to flexible cystoscopy in the diagnosis of UBC in hematuria patients. | This analysis shows the potential for a non-invasive test to be added to the diagnostic pathway for hematuria patients suspected of having UBC. If the DCRSHP is applied targeting hematuria patients at low risk of UBC, then it has the potential to be both effective and cost-effective. |
| 2006 | Svatek, R. S., et al. | Decision tree | Cost analysis to evaluate implementation of a widespread screening program using NMP22, a noninvasive, urinary, bladder tumor marker. | NMP22 can be a cost-efficient screening tool especially for high-risk target populations compared to currently used screening. More research (RCTs) is needed to assess the accuracy of NMP22 in detecting BCA in completely asymptomatic cohorts. |
| 2013 | van Kessel, K. E. M., et al. | Microsimulation | CEA to compare FGFR3 mutation analysis of voided urine samples compared to cystoscopy in the surveillance of patients treated for non-muscle invasive urothelial carcinoma. | Cystoscopy partly replaced by FGFR3 mutation analysis of urine is a safe and cost-effective surveillance strategy. |
| 2009 | Walker, K., et al. | Cohort-based | Characterize the main polymorphisms in both NAT2 and NAT1 in terms of their effect on enzyme activity and frequency in the population, and evaluate their impact on increasing the risk of bladder cancer. | Incorporating allele frequency data in different populations and their effects on acetylation into a Monte Carlo modeling framework led to a population distribution of NAT2 activity that was bimodal and associated with considerable variability in each population assessed. The ratio of the median to the first percentile of NAT2 activity ranged from 7 in Caucasians to 18 in the Chinese population. Polymorphisms in NAT1 are generally associated with relatively minor effects on acetylation function, with Monte Carlo analysis indicating less interindividual variability than seen in NAT2 analysis. |
| 2018 | Wang, Z., et al. | Cohort-based | Compare the efficacy of low dose (27 mg) vs full dose (81 mg) BCG immunotherapy for patients with intermediate and high-risk non-muscle invasive bladder cancer (NMIBC) after TURB. | A low-dose BCG may act slightly better than a full-dose BCG for patients with intermediate and high-risk of NMIBC. Need for high-quality studies to confirm this finding. |
| 2021 | Weiss, L. D., et al. | Compartmental model | Understand and describe the impact of regulatory loops on promoting CSC enrichment and consequent loss of therapy response due to the reduced susceptibility of CSCs to drugs. | Results from this study can help with improving BCA treatment by understanding the mechanism and suggest ways to overcome the phenomenon of CSC-based therapy resistance. |
| 2020 | Werneth, C. M., et al. | Compartmental model | Evaluate the effectiveness of MCM (Medical Countermeasures) for reducing cancer risk due to radiation using the NASA Space Radiation Cancer Risk. | The study numerically quantifies the benefits of MCM (e.g., aspirin and warfarin) in reducing cancer risk in terms of REID medians for astronauts embarking on a one-year deep space mission scenario. |
| 2020 | Wettstein, M.S., et al. | Microsimulation | CEA to find the required efficacy at which a NT (novel therapies: 3 distinct strategies i.e., systemic, low-intensity, and high-intensity intravesical) ) can compete with ERC in terms of (QALE) for treating BCG unresponsive NMIBC cases . In total, 24 different efficacy thresholds (including the recommendations) were investigated. | Results suggest increase of the thresholds for complete response rate and recurrence-free survival respectively to promote the development of clinically meaningful NTs. |
| 1993 | Wientjes, M. Guillaume, et al. | Compartmental model | Study the pharmacokinetics and pharmacodynamics of intravesical chemotherapy (MMC), predict drug exposure in tumors in the bladder wall, and correlate it with antitumor effect. | Simulations showed that changes in treatment parameters would affect the therapeutic outcome in the following rank order: dose (most importantly), residual volume, urine production, dosing volume, urine pH, dwell time. |
| 2022 | Williams, S. B., et al. | Cohort-based | Cost analysis of practices incorporating BLC with HAL for surveillance of NMIBC in the clinic setting. | From the office/clinic perspective results suggest that use of flexible BLC for the surveillance of NMIBC may be cost-effective. |

| Table 14. Objectives and conclusions of eligible simulation studies for bladder cancer. | | | | |
|---|---|---|---|---|
| Year | Author | Model Type | Objective | Conclusion |
| 2013 | Wong, K. A., et al. | Microsimulation | CEA to evaluate safety, tolerability and effectiveness of office-based OLA, compared to local anesthetic, and IC, for treating NMIBC in an elderly population with and without photodynamic diagnosis (PDD). | Results demonstrate the long-term cost-effectiveness of OLA of NMIBC, especially in the elderly. |
| 2009 | Yoon | Compartmental model | Develop an active programmable remote steering mechanism and an efficient motion sequence for bladder cancer detection and postoperative surveillance. | Preliminary experimental test results ensure that the motion sequencing program and the steering mechanism will efficiently move the image probe to scan the entire surface of the bladder. |
| 2012 | Yuan, Y., et al. | Compartmental model | Simulate results from a Phase I clinical trial to evaluate the effect of combined concurrent MMC chemotherapy with thermotherapy (deep regional heating using BSD-2000 Sigma-Ellipse applicator) for treating NMIBC. | Results suggest that prospectively planning patients using the effective thermal conductivity can potentially improve treatment efficacy (compared to manual operator adjustments) by lowering discomfort from reduced hot spots in normal tissue. |
| 2013 | Zhang, Y., et al. | Microsimulation | Compare current international guidelines (EAU and AUA) with alternative surveillance strategies for low-risk bladder cancer patients based on QALYs. | EUA guidelines seem more effective in terms of QALYs, with age and comorbidity to be key factors affecting the effectiveness of the surveillance strategy. |
